# A novel model to predict age of respiratory syncytial virus infection from birth timing in relation to RSV circulation

**DOI:** 10.1101/2025.04.01.25325040

**Authors:** Chris G. McKennan, Tebeb Gebretsadik, Steven M. Brunwasser, Michael Nodzenski, Daniel J. Jackson, James E. Gern, Pingsheng Wu, Tina V. Hartert

## Abstract

Respiratory syncytial virus (RSV) is a common respiratory virus that infects all children by age 2 to 3 years of age and causes the greatest morbidity at the extremes of life. Recent evidence suggests that early-life RSV infection, defined using active and passive surveillance with quantitative polymerase chain reaction- and serology-identified infection, is causal for childhood asthma. As such, identifying infants that are likely to be infected with RSV during this critical susceptibility window has important implications for determining who is most at risk for chronic respiratory sequelae like asthma. However, identifying the age of RSV infection is impractical in large populations, as not all infections are symptomatic, and measurement thus requires time- and cost-intensive surveillance. To address this, we developed the first probability model for age of first RSV infection. It uses an infant’s birthdate, demographic covariates, and publicly available RSV circulation data to determine the probability they were first infected at any age from birth to one year. Our model is easy to interpret, provides an exceptional fit for the data, and generalizes across populations, where we use it to accurately predict age of first infection in two independent cohorts. Our work represents a major development in RSV research, as it facilitates, for the first time, reliable estimation of the age of infant RSV infection during the first year of life in populations without the need for active surveillance.

## Introduction

Respiratory syncytial virus (RSV) is a common infant and childhood respiratory virus, with roughly half of all infants being infected by age one year and nearly all children by age three years (*1–3*). The age of first RSV infection has important implications not only for risk of severe disease but also for childhood wheeze and asthma risk. We have previously found that infants infected before age one year, with infection identified by quantitative polymerase chain reaction (PCR)- or serology-identified infection, are significantly more likely to develop asthma by age five years (*3*). We’ve also previously demonstrated that the relationship between date of birth in relationship to RSV circulation predicts asthma risk with children born around four months prior to the peak of RSV season at greatest risk of developing childhood asthma (*4*). Hence, identifying infants likely to be infected during this critical susceptibility period has important implications for determining those at the greatest risk of severe acute infection as well as developing chronic respiratory sequelae. However, identifying the age of first RSV infection is impractical in large cohorts because of the time and monetary costs, as not all infections are symptomatic and measurement thus requires active population surveillance.

An accurate *in silico* prediction of age of first RSV infection is an ideal alternative to costly surveillance. One possibility is to use existing models that predict RSV bronchiolitis events requiring healthcare visits (*4–6*). However, predicting age of RSV infection is very different from predicting RSV infection, as severe clinical manifestations of RSV occur among risk groups and constitute only a small fraction of all infections (*3, 7*). To address these limitations, we developed the first probability model for age of first RSV infection. Our model uses an infant birthdates, demographic information, and publicly available RSV circulation data from the Centers for Disease Control and Prevention (CDC) to determine probability of being infected for the first time at any age between birth and one year. The foundation of our model is the relationship between birthdate and the seasonal circulation and prevalence of RSV, which generally peaks in the winter in temperate climates with an absence of infections in the summer. For example, the probability an infant born in July is infected around six months will be large, as they will be six months in January when RSV circulation is at its peak.

In this study, we developed and tested our model in four independent cohorts using RSV surveillance data. In particular, we sought to determine how well our model (i) fits the observed data, (ii) predicts age of first RSV infection, and (iii) identifies age of first RSV infection without having to perform active RSV surveillance.

## Results

### Cohort demographics and RSV prevalence over time

Table 1 provides an overview of the study designs and demographic information for the four cohorts considered in this study. Birthdates were approximately uniform across the birth months considered in each cohort (Figure S1). Additional details regarding the cohorts used in this study have been previously reported (*8–11*).

**Table 1:** Study designs and demographic information for the four cohorts in our study. The total number of subjects in INSPIRE, COAST, and URECA is the number of subjects known to be infected or not infected with RSV by age one year. The total in PRIMA is the number of children with an asthma diagnosis at age four years.

|  | **INSPIRE**<br>(*n* = 1,741) | **COAST**<br>(*n* = 242) | **URECA**<br>(*n* = 301) | **PRIMA**<br>(*n* = 152,622) |
| --- | --- | --- | --- | --- |
| **Study design** |  |  |  |  |
| **Birth months** | June – December | January – December | January – December | January – December |
| **Study period**<br>(First birth – last birth + 1 year) | June 2012 – December 2014 | November 1998 – May 2001 | February 2005 – March 2008 | January 1995 – December 2004 |
| **Study location (%)** | Tennessee (100%) | Wisconsin (100%) | Baltimore, Maryland (28%)<br>New York, New York (13%)<br>Boston, Massachusetts (28%)<br>St. Louis, Missouri (31%) | Tennessee (100%) |
| **RSV serology at one year of age?** | Yes | Yes | Yes | No |
| **Surveillance during first year of life** | RSV qPCR following minimal infection symptoms | RSV qPCR following minimal infection symptoms | N/A | Bronchiolitis healthcare encounters |
| **Number of subjects with known age of first RSV infection (%)** | 361 (21%) | 53 (22%) | 0 (0%) | 0 (0%) |
| **Demographic data** |  |  |  |  |
| **Race/ethnicity: *n* (%)** |  |  |  |  |
| *Black* | 308 (18%) | 7 (3%) | 224 (74%) | 55,862 (37%) |
| *Hispanic* | 156 (9%) | 0 (0%) | 52 (17%) | 6,374 (4%) |
| *White* | 1,131 (65%) | 233 (96%) | 2 (1%) | 84,053 (55%) |
| *Other/Missing* | 146 (8%) | 2 (1%) | 23 (8%) | 6,333 (4%) |
| **Sex: *n* (%)** |  |  |  |  |
| *Female* | 828 (48%) | 103 (43%) | 151 (50%) | 73,930 (48%) |
| *Male* | 913 (52%) | 139 (57%) | 150 (50%) | 78,692 (52%) |
| **Daycare attendance in first year of life: *n* (%)** |  |  |  |  |
| *Yes* | 578 (33%) | 128 (53%) | 98 (33%) | 0 (0%) |
| *No* | 1,135 (65%) | 114 (47%) | 203 (67%) | 0 (0%) |
| *Missing* | 28 (2%) | 0 (0%) | 0 (0%) | 152,622 (100%) |
| **Older siblings: *n* (%)** |  |  |  |  |
| *Yes* | 876 (50%) | 132 (55%) | 67 (22%) | 90,136 (59%) |
| *No* | 865 (50%) | 110 (45%) | 232 (77%) | 62,373 (41%) |
| *Missing* | 0 (0%) | 0 (0%) | 2 (1%) | 113 (<0.1%) |
| **Maternal asthma: *n* (%)** |  |  |  |  |
| *Yes* | 342 (20%) | 100 (41%) | 135 (45%) | 3,884 (2.5%) |
| *No* | 1,398 (80%) | 142 (59%) | 165 (55%) | 83,264 (55%) |
| *Missing* | 0 (0%) | 0 (0%) | 1 (0.3%) | 65,474 (43%) |
| **Breastfeeding: *n* (%)** |  |  |  |  |
| *Ever breastfed* | 1,383 (79%) | 196 (81%) | 154 (51%) | 0 (0%) |
| *Never breastfed* | 345 (20%) | 46 (19%) | 142 (47%) | 0 (0%) |
| *Missing* | 13 (1%) | 0 (0%) | 5 (2%) | 152,622 (100%) |
| **Insurance: *n* (%)** |  |  |  |  |
| *Private* | 795 (46%) | 0 (0%) | 16 (5%) | 0 (0%) |
| *Public* | 923 (53%) | 0 (0%) | 277 (92%) | 152,622 (100%) |
| *Other/Missing* | 23 (1%) | 242 (100%) | 8 (3%) | 0 (0%) |

We determined the prevalence of RSV from January 1995 to April 2020 for all geographical regions in the United States (see Methods). Figure 1A contains an example for the region and time period covering INSPIRE participants. Figure 1B is the output of our smoothing and standardization pipeline, where the prevalence of RSV is assumed to be proportional to y-axis in Figure 1B.

**Figure 1.**
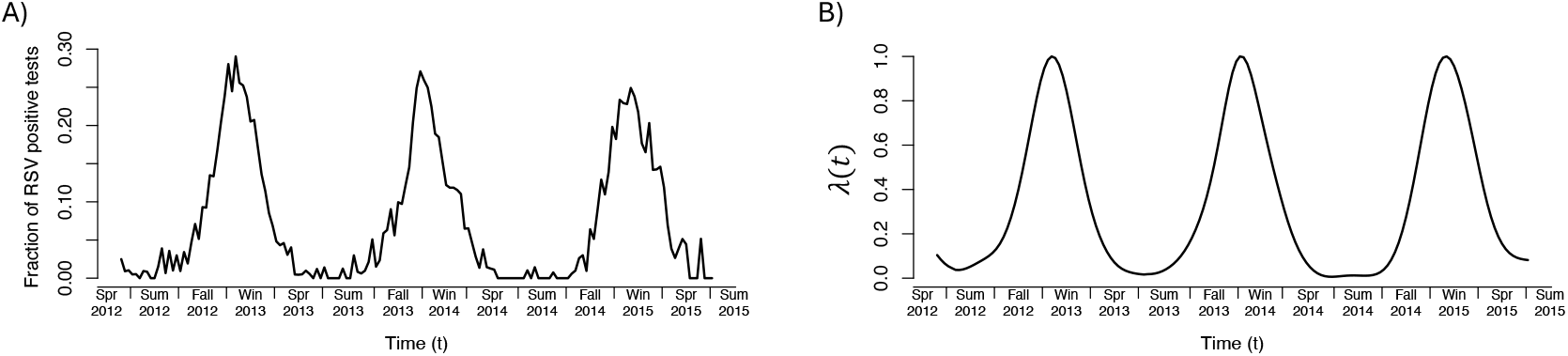
The prevalence of RSV over time from Spring 2012 to Summer 2015 in the geographical region covering INSPIRE participants. **A)** The raw data as reported by the CDC. **B)** The output of our smoothing and standardization pipeline applied to the raw data in A).

### Parameter estimates and model evaluation

Our model is based off a model for the instantaneous risk of first RSV infection, also called the “hazard function” in survival analysis. It assumes an infant’s risk of first being infected at age *a* equals the known prevalence of RSV at the time they are age *a* (which depends on their birthdate) times an unknown age-dependent weight function *w*(*a*), which captures age-related variation in RSV infection risk. Non-birthdate covariates are incorporated using a proportional hazards assumption (see Methods).

We used the INSPIRE cohort to estimate model parameters. Figure 2A shows that the weight function is small for ages close to zero, indicating newborns are less likely to be infected by RSV than older infants. The function peaks at around 6.5 months, suggesting this may be the time infants are at the greatest risk of infection. While it appears to decrease after eight months, the wide confidence intervals and dearth of infections during these months suggest the decrease during this time can not be confirmed given the lack of data. A sensitivity analysis indicated the shape of the weight function, e.g., its maxima at 2 months and 6.5 months and nadir at 4 months, was consistent when we re-estimated the weight function after partitioning INSPIRE subjects by race, sex, daycare attendance, or number of older siblings (Section S1.2).

**Figure 2.**
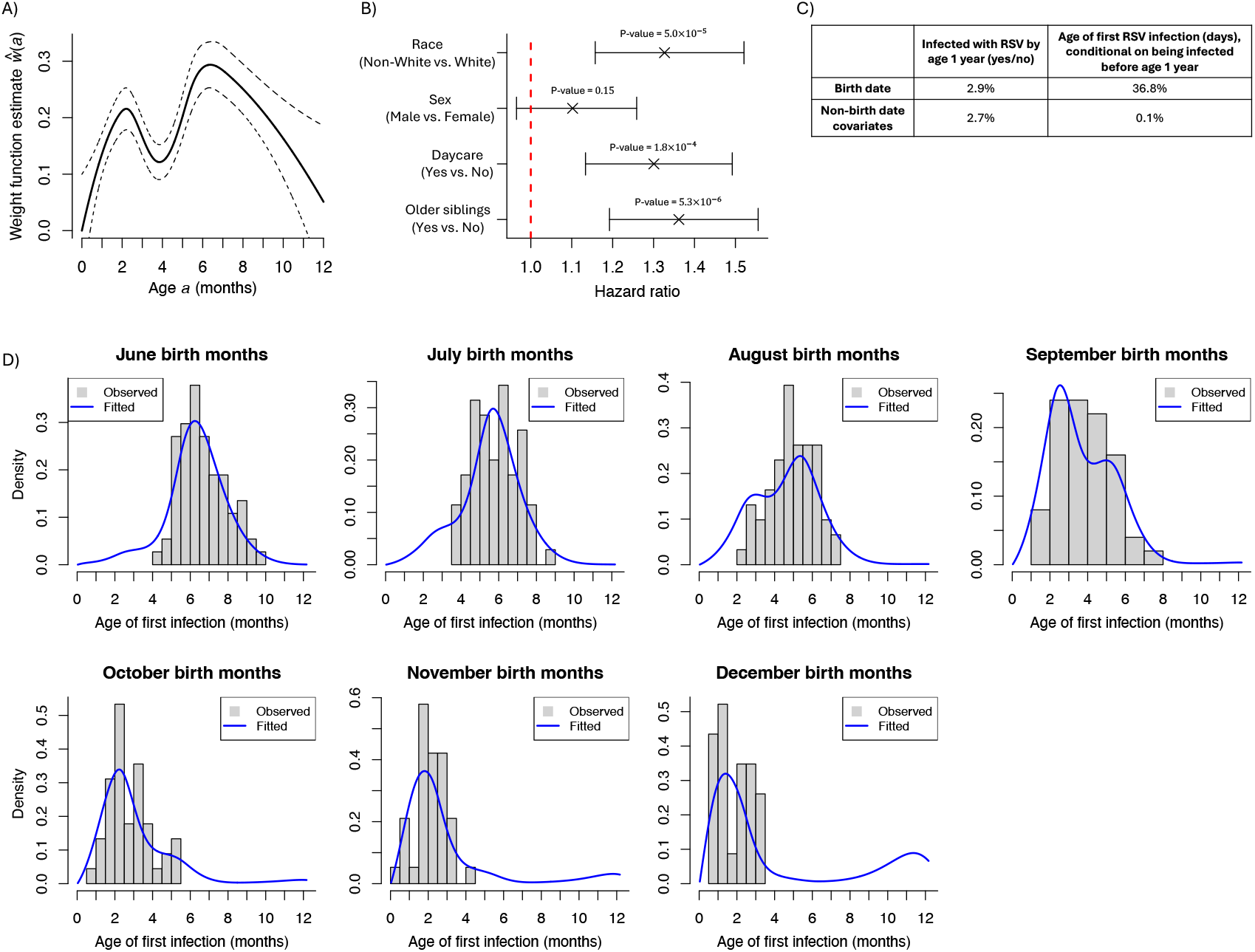
Estimates from INSPIRE. **A)** The estimated weight function (solid line) and 95% confidence intervals (dashed lines). **B)** Estimates and 95% confidence intervals for the effect of non-birthdate covariates. Estimates were exponentiated to get hazard ratios. P-values test the null hypothesis that the hazard ratio is 1. **C)** The percent of the variance of each outcome (columns) explained by each covariate (rows). **D)** The average estimated probability density function for INSPIRE subjects born in June through December. Probability densities were computed conditional on being infected before age one year.

Figure 2B gives the estimate for the effect of non-birthdate covariates expressed as a hazard ratio. As three of these were clearly important in predicting age of first infection, we sought to determine the relative importance of birthdate and the non-birthdate covariates we considered in predicting age of first infection. As Figure 2C indicates, the importance depends on the outcome one is trying to predict. If the goal is to try to predict whether an infant is infected by age one year, then birthdate and non-birthdate covariates have similar modest impacts. However, if it is known that an infant is infected by age one year and we would like to predict the exact age of first infection, birthdate is unequivocally more important and explains nearly 37% of the variance in infection age. The impact of birthdate on the outcome “infection by age one year” is dramatically smaller because all infants, regardless of birthdate, experience at least one RSV season during a one year interval. Section S2.7 details how we computed percent variance explained.

We next evaluated how well our model fit the observed data in INSPIRE. Figure 2D gives the estimated probability density functions for age of first infection for subjects born in June through December. By design, no INSPIRE subjects were born in January through May. These estimates are strikingly accurate, which is quite remarkable given that the effective number of parameters used to fit them was only 10.6 (6.6 parameters from *w*(*a*) and 4 parameters from non-birthdate covariates) (*12*).

### Testing our estimated model in two independent cohorts

To evaluate our model’s prediction accuracy, we used our estimates derived from INSPIRE to predict age of first infection in the COAST and URECA cohorts. Specifically, we sought to predict (i) which subjects were infected by age one year, as measured by RSV surveillance and serology, and (ii) the exact age of first infection for subjects with known age of first infection. For (i), we were particularly interested in assessing whether our model was well-calibrated. For example, if our model reports a subject was infected by age one year with probability 0.3, then 30% of all subjects with that infection probability should actually be infected by year one. Figure 3A gives the calibration plot and suggests our model is well-calibrated, as points lie on or near the dashed red line and 18 out of 20 points (90%) lie within the 95% confidence intervals (dashed grey lines). We combine COAST and URECA in Figure 3A to ensure probability bins had a sufficient number of subjects. Figure S4 gives separate calibration plots for COAST and URECA subjects.

**Figure 3.**
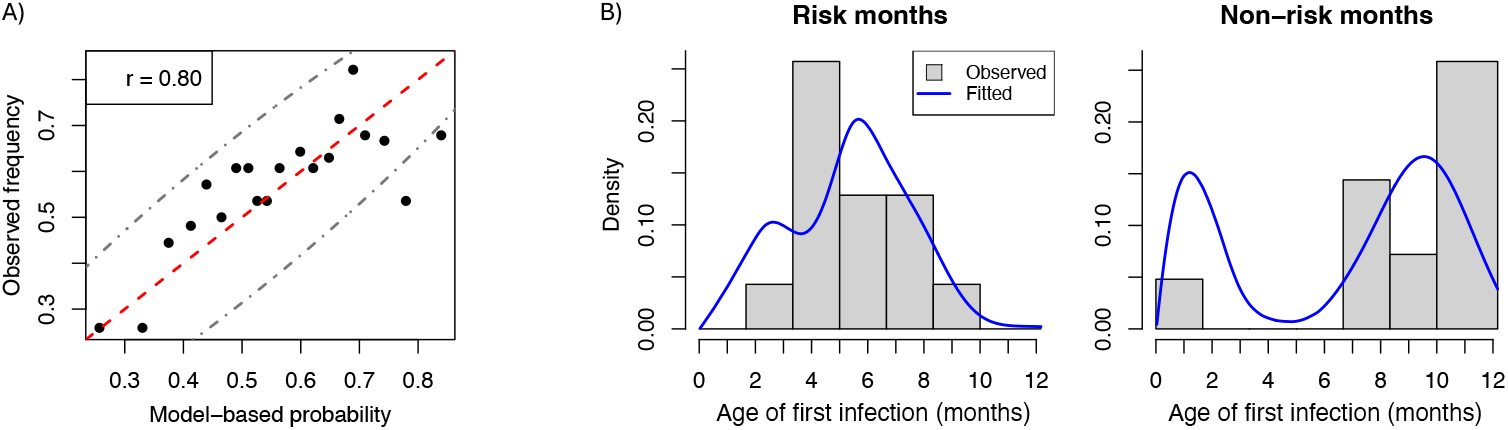
Predicting age of first RSV infection in COAST and URECA using parameter estimates obtained from INSPIRE. **A)** Model calibration curve. COAST and URECA subjects were binned by their model-based probability they were infected by age one year; bins contained between 27 and 28 subjects. The x-axis is the average probability in each bin and the y-axis fraction of subjects in each bin that were actually infected by age one year. The dashed red line is the line *y* = *x* and dot-dashed grey lines are 95% confidence intervals. **B)** The estimated probability density function for COAST subjects with a measured age of first infection before age one year that were born in risk months (*n* = 28) and non-risk months (*n* = 25). Fitted densities are conditional on being infected before age one year.

For (ii), we evaluated whether our probability density function estimate for age of first infection mirrored the distribution of observed first infection ages in the COAST cohort. URECA subjects did not have age of first infection data. We grouped COAST subjects by birth month, where we defined the “risk month” and “non-risk month” groups to be those born in June through December and January through May, respectively. We could not consider finer partitions of birth months because of sample size limitations in COAST. Figure 3B gives the probability density estimates in these two groups, which closely resemble the observed distributions.

### Using bronchiolitis healthcare visits to estimate model parameters

As there are few RSV surveillance studies, we lastly sought to determine whether bronchiolitis healthcare visits from the PRIMA cohort could be used to estimate the relationship between age of first RSV infection and birthdate. Due to the availability of birth and healthcare visit timing data, the weight function *w*(*a*) was estimated assuming it was a step function with steps (i.e., discontinuities) at each month (see Methods). We did not consider non-birthdate covariates because we found their impact in INSPIRE to be minor.

Figure 4A plots the estimated weight function. While it is similar to the estimate from INSPIRE, the uncertainty in the estimate from PRIMA is substantially larger, where one would need to increase the number of PRIMA study subjects from 1.5 *×* 10^5^ to 1.5 *×* 10^6^ to obtain standard errors equal to those from INSPIRE (see Section S1.3 for details). This considerable uncertainty stems from the fact that when using bronchiolitis healthcare visits, one must additionally estimate the nuisance function *c*(*a*), which is the probability an infection at age *a* becomes bronchiolitis requiring a healthcare visit (see Methods). As *c*(*a*) is highly non-uniform (see Section S1.3), it is difficult to distinguish between healthcare visits that are due to an increase in pathogen exposure (i.e., a change in *w*(*a*)) from those arising because subjects were more prone to bronchiolitis (i.e., a change in *c*(*a*)). Consequently, estimates for *w*(*a*) and *c*(*a*) were highly correlated.

**Figure 4.**
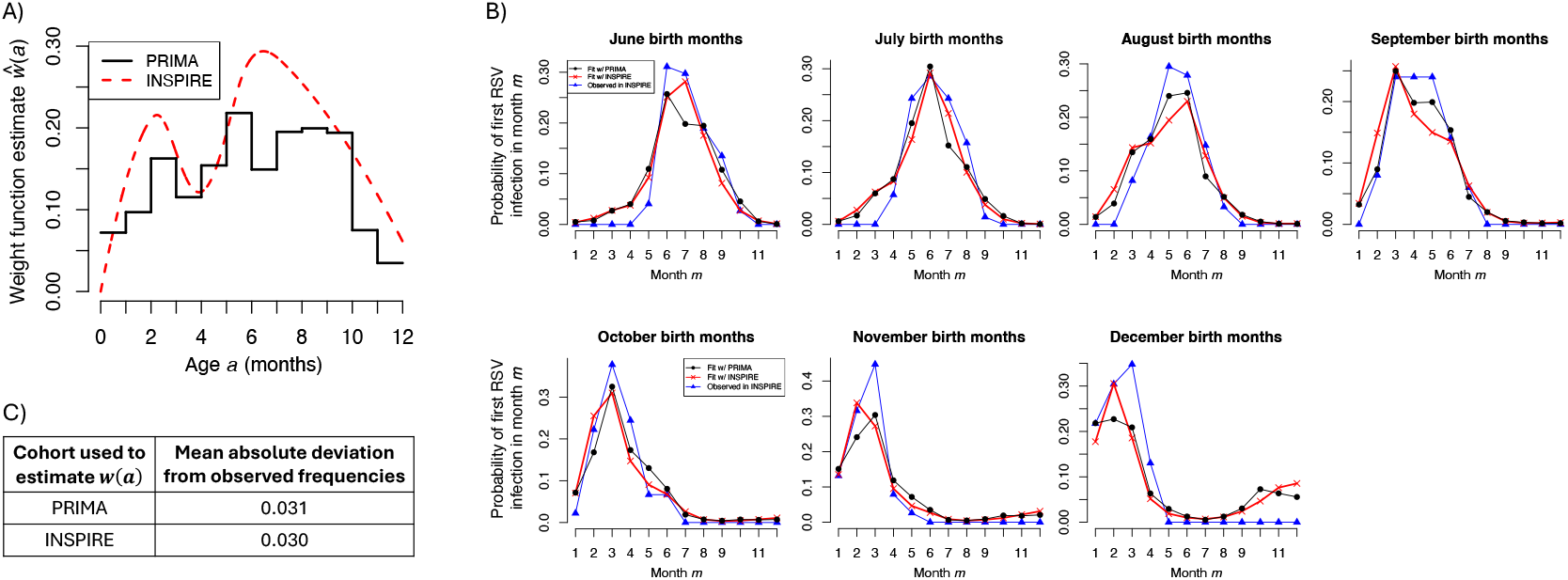
Estimates derived from PRIMA compared to those from INSPIRE. **A)** The weight function. **B)** The estimated probability subjects from INSPIRE were first infected with RSV in each month during the first year of life using the weight function estimated from PRIMA (black) or INSPIRE (red). The blue curves give the frequencies observed in INSPIRE. Probabilities are conditional on being infected in the first year of life. **C)** The mean absolute deviation between the estimated probabilities and observed frequencies in B).

Despite the increase in uncertainty, we evaluated whether our estimate for *w*(*a*) from PRIMA could be used to model age of first RSV infection in INSPIRE subjects. As PRIMA and INSPIRE are independent cohorts, this evaluation was equivalent to testing our model in an independent cohort. Figure 4B shows that model-based probabilities using the PRIMA-derived *w*(*a*) (black curves) closely resemble first infection frequencies observed in INSPIRE (blue curves). These probabilities were just as accurate as those obtained using the INSPIRE-derived *w*(*a*) (Figure 4C), which is quite remarkable given that INSPIRE-derived probabilities and observed frequencies were obtained from the same subjects.

## Discussion

In this study, we propose a novel probability model that utilizes publicly available RSV circulation data from the CDC to model age of first RSV infection as a function of birthdate and other demographic covariates. Our approach was to model the instantaneous risk of first infection, also known as the “hazard function” in the field of survival analysis. This was taken to be the product of the known prevalence of RSV at that time, which was determined by an infant’s birthdate and RSV surveillance data from the CDC, and an unknown age-dependent weight function. The weight function captures the age-dependent variation in RSV infection risk at a fixed point in time. Since we model hazard functions, we used the proportional hazards assumption to incorporate non-birthdate covariates. We could not use a standard Cox proportional hazard analysis to predict infection age because the proportional hazard assumption did not hold for the birthdate covariate.

We took a non-parametric approach to model the weight function and estimated it, along with the impact of non-birthdate covariates, using RSV surveillance data from the INSPIRE cohort. This resulted in an exceptional fit for the data, where our estimated model was able to almost perfectly capture the relationship between birthdate and first infection age. This is quite remarkable, as the effective number of parameters in our model was only 10.6 (*12*). Our estimates for the impact of non-birthdate covariates are consistent with those from a recent study examining the risk factors for RSV infection prior to age one year, as measured by serological testing (*2*).

We tested our estimated model in two independent cohorts, showing that our model-derived probabilities that subjects were infected by age one year were well-calibrated. We also showed that our estimates for the distributions of infection ages for infants known to be infected by age one year closely mirrored observed distributions. As the two testing cohorts were starkly different from INSPIRE in terms of their racial composition, geographic locations, study periods, and birth months (Table 1), these results indicate that our proposed model, and estimates for its parameters made in INSPIRE, generalize across populations.

The generalizability of our model estimated in INSPIRE, as well as our quantification of the importance of birthdate and non-birthdate covariates in predicting infection outcomes (Figure 2C), suggest a way to perform “RSV surveillance” in early life without actually having to conduct active surveillance to capture age of infection. Notably, if we know an infant was infected in the first year of life, birthdate alone explains nearly 37% of the variance in the age of infection. A potential RSV surveillance study design is therefore to perform serological testing on samples collected at age one year to determine whether the infant was infected with RSV prior to age one year, and use our model to predict age of infection. It is possible to forgo serological testing and use our model to also predict whether an infant was infected by age one year, although Figure 2C suggests this prediction may be noisy.

Since there are few RSV surveillance studies, we lastly tested whether we could use bronchiolitis healthcare encounters from the PRIMA cohort to estimate model parameters. This required making two concessions. First, we needed a set of assumptions, the most important being that the weight functions for RSV and the set of pathogens that also cause bronchiolitis and whose seasonality matched RSV’s were equal up to a constant of proportionality (see Methods). For example, if a six-month-old’s risk of infection with RSV was twice as large as a one-month-old’s, their risk of infection with the aforementioned pathogens was also twice as large. Second, we needed to estimate the nuisance function *c*(*a*) representing the probability an infection at age *a* turns into bronchiolitis requiring a healthcare encounter, leading to larger standard errors in the estimated weight function. Despite these concessions, the estimated weight function closely resembled the weight function estimated in INSPIRE, and we used it to almost perfectly recapitulate the observed distribution of infection ages in INSPIRE. These results suggest that while additional assumptions and a substantially larger sample size are needed to utilize bronchiolitis data, they can be used to estimate model parameters if surveillance data are limited or not available.

The weight function estimated in the INSPIRE and PRIMA cohorts in Figure 4A quantifies the age-dependent risk of RSV infection at a fixed point in time. It is close to zero for newborn infants, likely because newborns are both generally less exposed to the environment and may be protected from RSV by maternal antibodies (*13*). The peak at 6.5 months may be attributed to greater exposure to RSV at this age, and while maternal antibodies are not thought to prevent infection (they provide protection against severe disease), this period also coincides with the waning of maternal RSV antibodies (*13, 14*). The estimate in INSPIRE also exhibits a clear bimodal pattern, which was present even after re-estimating parameters in different subpopulations of INSPIRE. Bimodality is also evident in the estimate from PRIMA, which, taken together with the generalizability of estimates from INSPIRE to other populations, suggests the pattern is not a statistical anomaly or an artifact of cohort design. The decrease from the first mode at two months to the nadir at four months could reflect an improved T cell-mediated immune response, as this coincides with the time that naïve CD4+ T cells are highest in infants (*15*), which confer robust protection from lung infections in infancy (*16*).

Despite our model’s exceptional performance, we must acknowledge a few limitations. First, due to the availability of surveillance data in INSPIRE, we modeled age of first RSV infection up to age one year. While it may be possible to extend the model past one year, care would have to be taken when curating surveillance data to avoid mislabeling a second RSV infection in year two as a first infection. We circumvented this issue because so-called “repeat infections” before age one year are rare and, when they do occur, are infections from the same virus, likely representing infections that were not cleared by the host (*17*). It is also likely that infection before age one year has the greatest implication for childhood asthma-related disorders (*3, 4*). Second, we standardized RSV seasonality so that the maximum prevalence was the same across RSV seasons (Figure 1B), which helped avoid biases caused by year-to-year variation in RSV testing frequency (*18*). This implicitly assumes that the amount of circulating RSV is constant across seasons, which is reasonable given that the limited number of surveillance studies all estimate that about half of all infants are infected by age one year (*1–3*). Further, our estimated model generalized to cohorts with subjects sampled at different times and from different locations, suggesting the impact of seasonal variation in circulating RSV is trivial. Lastly, the timing of completely asymptomatic infections detected only by surveillance were not available in INSPIRE, suggesting our estimated model in INSPIRE may be more reflective of symptomatic infections. However, several features of our experimental design and results indicate this is not the case. First, minimal symptoms were required to trigger a nasal collection for PCR in INSPIRE and were not visits that required a healthcare encounter (*3*). Second, our model was able to predict infection in two independent cohorts, which included both symptomatic and asymptomatic infections. Third, estimates in INSPIRE closely mirrored those from PRIMA, where our analysis in PRIMA accounted for symptomatology by modeling the probability an infection turned into a symptomatic infection requiring a healthcare visit.

In summary, we have developed, to our knowledge, the first probability model to predict age of first RSV infection. Our model utilizes publicly available RSV circulation data from the CDC and a subject’s birthdate, as well as other other demographic covariates, to provide personalized RSV infection risk prediction. Our model is interpretable, yields an exceptional fit to the data, generalizes across populations, and can even be fit using easy-to-obtain bronchiolitis healthcare encounter data. We believe our work represents a substantial development in viral surveillance, as it allows, for the first time, identification of age of infant RSV infection within large populations without the need for active surveillance.

## Materials and Methods

### Study design

We included data from four independent cohorts: Infant Susceptibility to Pulmonary Infections and Asthma Following RSV Exposure (INSPIRE), Childhood Origins of Asthma (COAST), Urban Environment and Childhood Asthma (URECA), and Prevention of RSV: Impact on Morbidity and Asthma (PRIMA). In our primary analysis, we used RSV surveillance data available in INSPIRE to estimate model parameters and tested the performance of our estimated model in COAST and URECA. We used bronchiolitis healthcare visits available in the PRIMA cohort to determine whether model parameters could also be estimated from easy-to-obtain healthcare visit data.

### Age of first RSV infection measured in the INSPIRE, COAST, and URECA cohorts

The age of first RSV infection was measured for subjects participating in the INSPIRE study, our discovery cohort, during their first year of life. Participants were born between June and December (*10*). Briefly in INSPIRE, active and passive surveillance was conducted during infancy, with every two week assessment of respiratory symptoms, and nasal sample run for RSV PCR for those with symptoms. Since not all RSV infections manifest symptoms, all infants had RSV serology measured at age one year to determine which infants were infected by age one year (*3*).

Age of first infection in the COAST cohort, our validation cohort, was measured similarly (*8*, 19, 20). Subjects missing an infection age were assigned an infection status (infected or not infected) using the same serological testing as in INSPIRE. In the URECA cohort only one-year of age RSV serology data were available.

### Bronchiolitis healthcare visits in the PRIMA study

We used bronchiolitis healthcare visits from infants ≤ 1 year of age that were a part of the PRIMA study to determine whether we could estimate model parameters from easy-to-obtain healthcare visits. We considered PRIMA subjects that were enrolled in Tennessee Medicaid Program and were born prior to 2004 so that an asthma diagnosis at age four years would be available (*21*). Healthcare visits were hospitalizations, emergency room visits, or physician office visits. Infants’ age at the first bronchiolitis healthcare visit was determined.

### Determining RSV prevalence over time

We used publicly available data from the CDC to determine *λ*(*t*), the the prevalence of RSV over time *t*, for 17 different geographical regions in the United States defined by Health and Human Services (HHS) regional offices (1-10) and Census Bureau divisions (East North Central, West North Central, East South Central, West South Central, Mid-Atlantic, South Atlantic, New England, Mountain, Pacific). While *λ*(*t*) is region-specific, we suppress its dependence on region to avoid excess notation. Since the below model only depends on knowing *λ*(*t*) up to a constant of proportionality, we define *λ*(*t*) here up to a multiplicative constant. That is, *λ*(*t*) actually gives the relative prevalence of RSV, where the relative amount of circulating RSV in a geographical region at any times *t*_1_ and *t*_2_ is *λ*(*t*_1_)*/λ*(*t*_2_).

The data for each geographical region consisted of the number of viral samples and the fraction of them that were RSV positive, determined by culture, antigen, or PCR testing. Data were reported on a weekly basis starting in January 1995. We determined the fraction of positive tests over time using antigen testing because antigen tests were reported in the greatest number over the course of our study; PCR test results were not available prior to 2004 (Figure S5). We defined *λ*(*t*) to be the fraction of positive tests after smoothing and standardizing so that the maximum height of each peak was one (Figure 1; see Section S2.1 for details). Standardizing accounted for the year-to-year variation in the frequency of RSV testing (*18*).

We mapped study locations in each cohort (Table 1) to geographical regions by determining the HHS office and Census Bureau division that covered them.

### Probability model for age of first RSV infection

Here we describe our model relating a subject’s birthdate to their age of first RSV infection. We show how to incorporate additional covariates below. Let *i* index a subject, *B*_*i*_ be their birthdate, and *R*_*i*_ be their age of first RSV infection. We modeled age of first infection by modeling the instantaneous risk of first infection, which is defined as the likelihood a subject is infected at age *a* given they have not been infected up until that point. This is also known as the “hazard function” in survival analysis.

Recall *λ*(*t*) is proportional to the prevalence of RSV at time *t*. Since infection risk ought to be proportional to the amount of RSV in the population, a naïve model would be to assume that instantaneous risk is proportional to RSV prevalence:

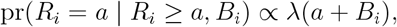

where *a* + *B*_*i*_ is the time at which subject *i* is age *a*. However, this model ignores the fact that one’s infection risk also depends on age, as very young infants are likely less exposed to RSV (and the environment in general) and have more maternally derived RSV antibodies than older infants (*1*). We therefore modified the above model to assume instantaneous risk depends on both RSV prevalence and age:

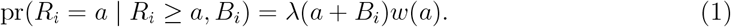

Here, *w*(*a*) is a non-negative weight function that captures the age-dependent risk of being infected by RSV. Since very young infants have the least exposure to RSV and greatest levels of maternally derived RSV antibodies, and exposure increases and maternally derived antibodies decrease over time (*1*), we expect *w*(*a*) to be small when age *a* is close to zero and get larger as *a* increases.

The instantaneous risk model in Equation 1 completely determines the probability model relating age of first infection and birthdate, as it implies the likelihood for subject *i* can be expressed as

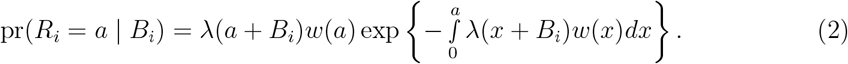

### Estimating the weight function

Since the function *λ*(*·*) is known, the only unknown in the likelihood given by Equation 2 is the age-dependent weight function *w*(*a*). We assume *w*(*a*) is a continuous and smooth function and require *w*(*a*) ≥ 0 to ensure the likelihood is non-negative. As the functional form of *w*(*a*) is unknown, we parameterize *w*(*a*) using a cubic B-spline basis (*22*). Since the B-spline basis elements are themselves non-negative, we encode the non-negativity constraint in *w*(*a*) by requiring basis coefficients be non-negative.

We fit *w*(*a*) using data from the INSPIRE cohort, which contains data for subjects with an observed age of first infection *R*_*i*_ between zero and one year (*n* = 361), subjects known to be infected before age one year but without an observed *R*_*i*_ (*n* = 583), and subjects known to have their first infection after age one year but without an observed *R*_*i*_ (*n* = 797). Our estimator for *w*(*a*) is a penalized maximum likelihood estimator, where the penalty encourages *w*(*a*) to be smooth (i.e., a small second derivative). We set the number of knots in our B-spline parametrization of *w*(*a*) to be 18 (10 internal knots and eight boundary knots), which provided a sufficiently flexible model. While this ostensibly begets a large number of model parameters, the penalty meant the effective number of parameters used to model *w*(*a*) was 6.6 (*12*), implying over-fitting was not an issue. Using a smaller number of knots resulted in a nearly identical estimate (Figure S6). Sections S2.2-S2.3 provide additional details.

### Incorporating additional covariates into the model

Since our model from Equation 1 is a model on the instantaneous risk, otherwise known as the hazard function, we incorporate non-birthdate covariates using a proportional hazards assumption. That is, if *z*_*i*_ is a vector of non-birthdate covariates for subject *i*, we modify the instantaneous risk from Equation 1 to be

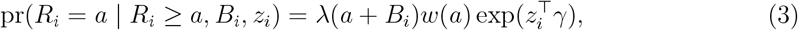

implying the likelihood from Equation 2 becomes

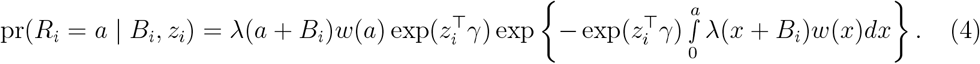

We detail how we jointly estimate *w*(*a*) and *γ* in Section S2.3. We included the covariates sex (male/female), race (White/non-White), daycare attendance (yes/no), and older siblings (yes/no) in our analysis of INSPIRE data. We did not include early-life nutrition (e.g., breast milk or not) because much of an infant’s protection to RSV derives from transplacental transfer of maternal antibodies (*13, 23*), and because over 80% INSPIRE subjects were breastfed.

### Testing the estimated model in the COAST and URECA cohorts

We used our INSPIRE-derived estimates for *w*(*a*) and *γ* in Equation 4 to estimate, for each COAST and URECA participant, (i) the probability they were infected with RSV by age one year and (ii) their probability density (Equation 4), which gives the likelihood they were first infected at age *a* for all *a* from 0 to 1 year. We included the same non-birthdate covariates as we did when fitting the model in INSPIRE (see above). Section S2.4 contains additional details.

### Estimating parameters using bronchiolitis healthcare visit data

We used the bronchiolitis healthcare visit data from the PRIMA cohort to determine whether we could use easy-to-obtain healthcare encounter data to estimate the weight function *w*(*a*). We ignored additional demographic covariates because we found their impact in INSPIRE to be minor (see Results).

It was not known whether bronchiolitis events in PRIMA were caused by RSV. While the seasonal prevalence of RSV and frequency of bronchiolitis health care visits are congruent (*4*), and most bronchiolitis health care visits during infancy are caused by RSV (*24–26*), it is possible that other pathogens whose seasonality overlaps with RSV circulation could cause bronchiolitis. To address this, we assumed that the age-dependent variation in risk of being infected by these pathogens was the same as it was for RSV. That is, the weight functions for these pathogens and RSV were the same up to a constant of proportionality. For example, if a six-month-old was twice as likely to be infected by RSV as a one-month-old (i.e., *w*(6 months) = 2*w*(1 month)), the risk of infection by these pathogens is also twice as large.

We additionally assumed that (i) each subject can be infected with RSV or the above mentioned pathogens at most once during the first year of life and (ii) the probability an infection turns into bronchiolitis requiring a healthcare visit depends on the infant’s age and not the date of the infection. Assumption (i) is motivated by the observation that so-called “repeat RSV infections” during the first year of life are rare and, when they do occur, have been shown to represent the same virus, presumably because it has not been cleared by the host (*17*). We assume (ii) because we know of no data suggesting otherwise.

Let *L*_*i*_ be the the age of the *i*-th subject’s first bronchiolitis healthcare visit and 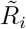 be the age of their first RSV infection or their first infection with one of the above mentioned pathogens whose seasonality overlaps with RSV circulation. Under the above assumptions, the likelihood for *L*_*i*_ conditional on birthdate *B*_*i*_ is

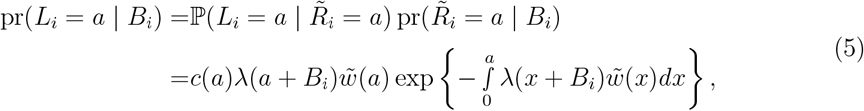

where 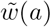 is proportional to *w*(*a*) and *c*(*a*) is unknown and is the probability an infection at age *a* turns into bronchiolitis. As we did in INSPIRE, we estimated 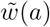, as well as *c*(*a*), up to age one year. Since only the years and months of birth and bronchiolitis healthcare visit dates were available in PRIMA, we assumed 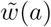 and *c*(*a*) were step functions with steps (i.e., discontinuities) at months one through 11. Since 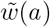 is proportional to *w*(*a*), we set our estimate for *w*(*a*) to be a normalizing constant times our estimate for 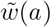, where the normalizing constant was chosen so that the resulting model-based probability an infant was infected with RSV before age one year matched the frequency observed in INSPIRE. Section S2.5 contains additional details.

We used our PRIMA-derived estimate for *w*(*a*) to estimate the probability INSPIRE subjects were first infected with RSV in each of their first 12 months of life. The mathematical expression is given in Section S2.6. We consider this probability and not the density like we did for COAST and URECA subjects because *w*(*a*) could only be estimated at the resolution of months in PRIMA.

## Supporting information

Supplementary results and methods; Supplementary figures S1-S8

## Data Availability

The RSV data were obtained from the National Respiratory and Enteric Virus Surveillance System (NREVSS). Readers can request the data from NREVSS at.

## Acknowledgements

We are deeply grateful to all of the families and children who participated in this study, to the INSPIRE research study staff, to the middle Tennessee pediatric practices with whom we collaborated to enroll a representative population of our region, to the Division of TennCare in the Tennessee Department of Finance and Administration, and the Tennessee Department of Health, Office of Policy, Planning & Assessment.

## Funding

NIH grants U19AI095227, UG3OD023282, UL1TR000445, 1U2410769079. The content is solely the responsibility of the authors and does not necessarily represent the official views of the funding agencies.

## Author contributions

Conceptualization: CGM, TG, PW, TVH

Methodology: CGM, TG, PW, TVH

Investigation: CGM, TG, SMB, PW, TVH

Visualizations: CGM, TVH

Funding acquisition: DJJ, JEG, PW, TVH

Writing: CGM

Writing – review & editing: CGM, TG, PW, SMB, MN, DJJ, JEG, PW, TVH

## Competing interests

CGM reports grants from NIH during the conduct of the study and personal fees from SignatureDx outside the submitted work. TVH is a member of NIH/NHLBI Council, the Parker B. Francis Family Foundation council of scientific advisors as a grant reviewer, serves as the co-chair of the ATS Vaccines and Immunization Initiative, content writer for UpToDate, and as a member of the RSV vaccine program DSMB for Pfizer. JEG is a consultant and has stock options in Meissa Vaccines Inc.

## Notes

### Funding Statement

This study was funded by NIH grants U19AI095227, UG3OD023282, UL1TR000445, 1U2410769079. The content is solely the responsibility of the authors and does not necessarily represent the official views of the funding agencies.

