## Supplementary results and methods; Supplementary figures S1-S8 for "A novel model to predict age of respiratory syncytial virus infection from birth timing in relation to RSV circulation"

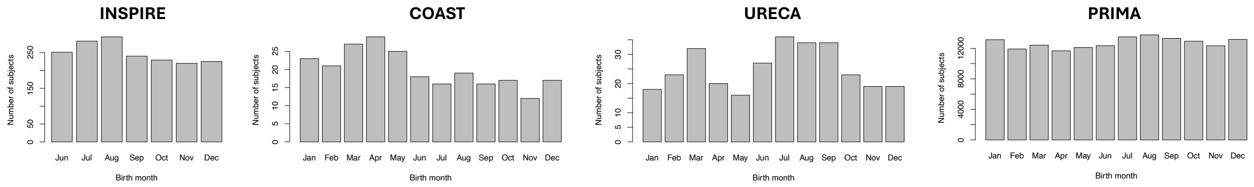


Figure S1: Birth month distributions in all four cohorts.

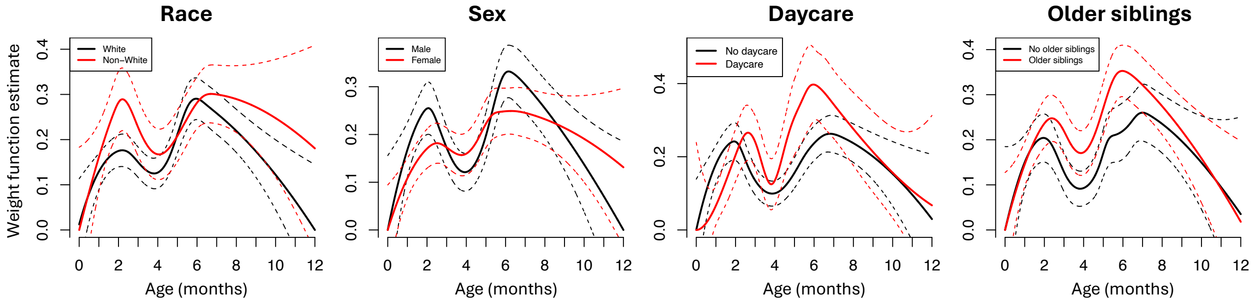


Figure S2: Estimates for the weight function in each subpopulation in INSPIRE (solid lines). Dashed lines give 95% confidence intervals.

### S1 Supplemental results

#### S1.1 Birth months

Figure S1 gives the birth months in all four cohorts.

#### S1.2 Additional results from INSPIRE

We performed a sensitivity analysis to check whether the shape of the weight function in Figure 2A was consistent across subpopulations in INSPIRE. Specifically, we split INSPIRE subjects by race (Non-White or White), sex (Male or Female), daycare attendance during the first year of life (Yes or No), or older siblings (Yes or No). Figure S2 shows that the shape of the weight function estimate is consistent across all of these subpopulations.

#### S1.3 Additional results from the PRIMA cohort

The calculation to determine we needed 10 times as many PRIMA subjects to obtain a weight function estimate as accurate as INSPIRE's was based off the standard error at each estimated weight function's maximum.

Figure S3 gives the estimate for $c(a)$. The standard errors for the estimate on the interval (10 months, 11 months] and (11 months, 12 months] were exceedingly large (1.1 and 4.8, respectively), suggesting the large estimates on these intervals is likely just due to estimation error. Even if we were to exclude these two noisy intervals, the estimate for $c(a)$ still looks highly non-uniform. To explicitly test the uniformity of $c(a)$, we tested the null hypothesis that $c(a)$ was a constant function. The p-value was $2.0 \times 10^{-7}$, indicating we should unequivocally reject the null.

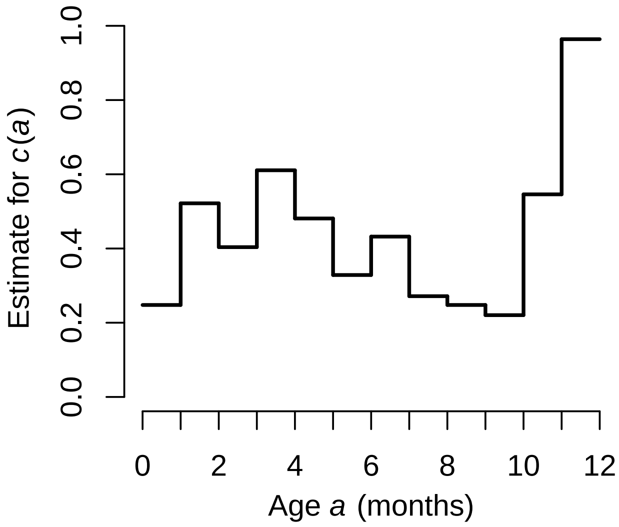


Figure S3: Estimate for $c(a)$ from the PRIMA cohort. The maximum value is 0.96.

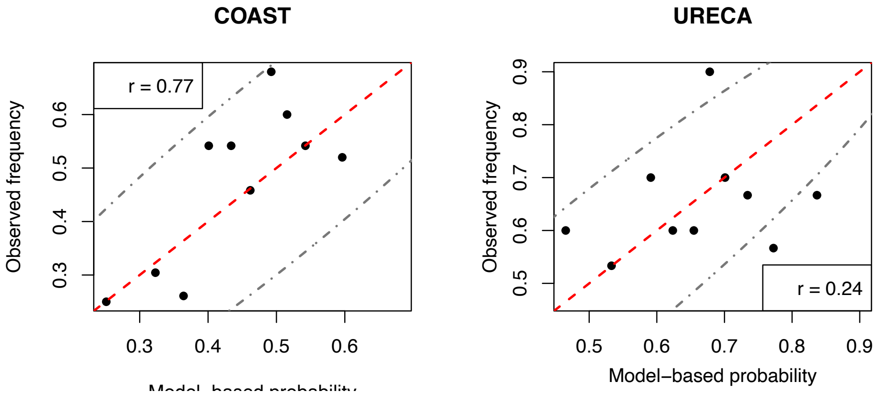


Figure S4: The analogue of Figure 3A, but plotted separately for subjects from the COAST and URECA cohorts. There were 23-25 and 29-30 subjects in each bin in the COAST and URECA plots, respectively.

##### S1.4 Additional results from COAST and URECA

Figure S4 gives the analogue of Figure 3A, but plotted separately for subjects from the COAST and URECA cohorts.

#### S2 Supplemental statistical methods

##### S2.1 Smoothing and standardization pipeline

Figure S5 gives the number of tests available in the geographic region covering PRIMA and INSPIRE subjects.

We smoothed the observed fraction of RSV positive tests to obtain $\lambda(t)$ using a three step procedure. Let $y_t$ and $n_t$ be the number of RSV positive tests and the total number of tests

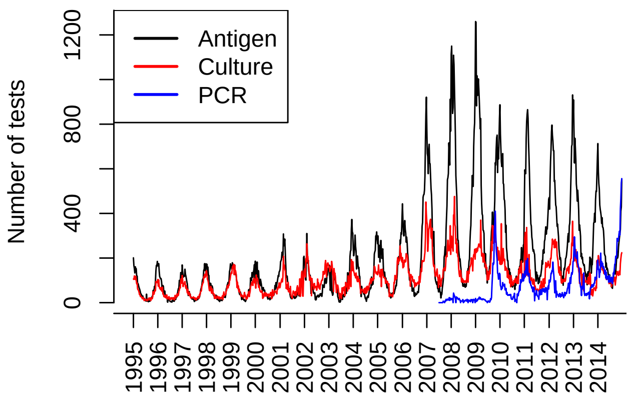


Figure S5: Number of tests sent to Health and Human Services (HHS) region 4 (the HHS region covering INSPIRE subjects).

at time $t$, where $t$ is at the resolution of weeks. We first obtained an estimator $\hat{p}_t^{(1)}$ for the true probability of a positive tests using local-linear Kernel smoothing:

$$\{\hat{\mu}_t^{(1)}, \hat{\beta}_t^{(1)}\} = \arg\max_{\mu_t, \beta_t \in \mathbb{R}} \sum_{t'} K\{(t-t')/h\} \log\{\text{Binom}(y_{t'}; \text{expit}(\mu_t + \beta_t(t'-t)), n_{t'})\}$$
$$\hat{p}_t^{(1)} = \text{expit}\{\hat{\mu}_t^{(1)}\}$$

Here, $\text{Binom}(y; p, n)$ is the likelihood of a binomial distribution with probability $p$ and number of trials $n$ evaluated at $y$, and $K(u) = (1-u^2)^2 1\{|u| \leq 1\}$ is the biweight kernel. We set the bandwidth $h$ to be 5 weeks. Note that this down-weights time points $t'$ where $n_{t'}$ is small.

We then smoothed $\hat{p}_t^{(1)}$ further using Gaussian smoothing:

$$\hat{p}_t^{(2)} = \sum_{t':|t-t'| \leq 2.5\sigma} \hat{p}_{t'}^{(1)} \phi_\sigma(t-t'),$$

where $\phi_\sigma(u)$ is probability density function of a normal random variable with mean zero and standard deviation $\sigma$. We set $\sigma$ to be 3 weeks, which implied $\hat{p}_t^{(2)}$ was smoothed using 15 weeks worth of data, which is about the length of one season.

The last step was to normalize $\hat{p}_t^{(2)}$. We defined each year to start July 1 and scaled $\hat{p}_t^{(2)}$ separately within each year so that the maximum value was 1. We called the resulting function $\lambda(t)$. We used linear interpolation to define $\lambda(t)$ at the resolution of days, and assumed $\lambda(t)$ was constant between days. That is, $\lambda(t)$ was a step function with discontinuities (steps) at each day.

#### S2.2 Estimating the weight function without non-birth date covariates

Here we describe how we estimate the weight function $w(a)$ when we exclude non-birth date covariates. Let $i$ index subject, $B_i$ be birth date, and $R_i$ be the age of the $i$-th subject's first RSV infection. We parameterize $w(a)$ using a cubic B-spline basis. To explicitly indicate the weight function's dependence on the unknown basis coefficients, we let $w(a) = w(a;\beta) = \sum_{j=1}^{J} \beta_j b_j(a)$, where $b_j(a)$ are the known cubic B-spline basis functions and $\beta_j$ are the unknown basis coefficients. Since $b_j(a)$ are themselves non-negative, we encode the non-negativity constraint in $w(a;\beta)$ by requiring the elements of $\beta$ be non-negative.

Define the index sets

$$\begin{aligned}\mathcal{I}_1 &= \{i : R_i \text{ is observed}\}\\ \mathcal{I}_2 &= \{i : R_i \text{ is missing and } R_i \leq 1 \text{ year}\}\\ \mathcal{I}_3 &= \{i : R_i \text{ is missing and } R_i > 1 \text{ year}\}.\end{aligned}$$

Equation 2 implies the log-likelihood can be expressed as

$$\begin{aligned}\ell(\beta) =& \sum_{i\in\mathcal{I}_1} \log\{w(R_i;\beta)\} - \int_0^{R_i} \lambda(x+B_i)w(x;\beta)dx\\ &+ \sum_{i\in\mathcal{I}_2} \log\left[1-\exp\left\{-\int_0^{1\text{ year}} \lambda(x+B_i)w(x;\beta)dx\right\}\right] - \sum_{i\in\mathcal{I}_3} \int_0^{1\text{ year}} \lambda(x+B_i)w(x;\beta)dx.\end{aligned}$$

The date $R_i$, when observed, is given at the granularity of days. And since $\lambda(t)$ is defined to be a step function with discontinuities at each day (Section S2.1), integrals in the above log-likelihood can be expressed as

$$\begin{aligned}\int_0^{R_i} \lambda(x+B_i)w(x;\beta)dx =& \sum_{j=1}^{J} \beta_j \int_0^{R_i} \lambda(x+B_i)b_j(x)dx = \sum_{j=1}^{J} \beta_j \sum_{a=1}^{R_i} \lambda(a+B_i) \int_{a-1}^{a} b_j(x)dx\\ =& \beta^\top v_i\end{aligned}$$

where $v_i$ is length-$J$ vector whose $j$-th element is equal to

$$\sum_{a=1}^{R_i} \lambda(a+B_i) \int_{a-1}^{a} b_j(x)dx.$$

Similarly,

$$\int_0^{1\text{ year}} \lambda(x+B_i)w(x;\beta)dx = \int_0^{365} \lambda(x+B_i)w(x;\beta)dx = \beta^\top v_i,$$

where the $j$-th element of $v_i$ is

$$\sum_{a=1}^{365} \lambda(a+B_i) \int_{a-1}^{a} b_j(x)dx.$$

As such, the log-likelihood can be expressed as

$$\ell(\beta) = \sum_{i\in\mathcal{I}_1} \log\{\beta^\top b(R_i)\} - \beta^\top v_i + \sum_{i\in\mathcal{I}_2} \log\{1-\exp(-\beta^\top v_i)\} - \sum_{i\in\mathcal{I}_3} \beta^\top v_i, \tag{S1}$$

where $b(a) = (b_1(a), \ldots, b_J(a))^\top$. It is easy to check that $\ell(\beta)$ is a concave function.

We define our estimator for $\beta$ to be the penalized maximum likelihood estimator:

$$\hat{\beta} = \underset{\substack{\beta \in \mathbb{R}^J \\ \beta \geq 0}}{\arg\max} \{\ell(\beta) - \alpha P(\beta)\} = \underset{\substack{\beta \in \mathbb{R}^J \\ \beta \geq 0}}{\arg\min} \{-\ell(\beta) + \alpha P(\beta)\}, \quad \alpha \geq 0$$

$$P(\beta) = \frac{1}{2} \sum_{j=3}^{J} (\beta_j - 2\beta_{j-1} + \beta_{j-2})^2 = \frac{1}{2} \beta^\top M \beta,$$

where $M$ is a known matrix encoding the penalty. The penalty function $P(\beta)$ resembles the integral of the squared second derivative of $w(a;\beta)$ and encourages $w(a;\beta)$ to be smooth [1]. As the penalty and minus log-likelihood are convex functions of $\beta$, we solve the above optimization using constrained Newton-Raphson updates. We choose $\alpha$ via 10-fold cross validation.

#### S2.3 Jointly estimating $w(a)$ and $\gamma$

When we incorporate non-birth date covariates $z_i \in \mathbb{R}^p$, the model from Equation 4 implies the log-likelihood in Equation S1 should be modified to be

$$\begin{aligned} \ell(\beta, \gamma) = & \sum_{i \in \mathcal{I}_1} \log\{\beta^\top b(R_i)\} + \gamma^\top z_i - \exp(\gamma^\top z_i) \beta^\top v_i \\ & + \sum_{i \in \mathcal{I}_2} \log \left[1 - \exp\left\{-\exp(\gamma^\top z_i) \beta^\top v_i\right\}\right] - \sum_{i \in \mathcal{I}_3} \exp(\gamma^\top z_i) \beta^\top v_i. \end{aligned}$$

We jointly estimate $\beta$ and $\gamma$ by solving an analogous penalized likelihood problem:

$$(\hat{\beta}, \hat{\gamma}) = \underset{\substack{\beta \in \mathbb{R}^J, \gamma \in \mathbb{R}^p \\ \beta \geq 0}}{\arg\max} \{\ell(\beta, \gamma) - \alpha P(\beta)\} = \underset{\substack{\beta \in \mathbb{R}^J, \gamma \in \mathbb{R}^p \\ \beta \geq 0}}{\arg\min} \{-\ell(\beta, \gamma) + \alpha P(\beta)\}, \quad \alpha \geq 0.$$

We solve this problem by iteratively minimizing the penalized minus log-likelihood over $\beta$ while fixing $\gamma$, which we do using the optimization from Section S2.2, and over $\gamma$ while fixing $\beta$, which we do using BFGS [2].

To obtain estimates for the variance of $\hat{\theta} = (\hat{\beta}^\top, \hat{\gamma}^\top)^\top$, we note that a Taylor expansion of the gradient of the penalized log-likelihood at $\hat{\theta}$ around the true parameter implies the variance can be approximated as

$$\mathrm{Var}(\hat{\theta}) \approx \{\nabla_\theta^2 \ell(\hat{\theta}) + \alpha M\}^{-1} \mathrm{Var}\{\nabla_\theta \ell(\theta^*)\} \{\nabla_\theta^2 \ell(\hat{\theta}) + \alpha M\}^{-1},$$

where $M$ is as defined in Section S2.2 and $\theta^*$ is the true parameter. As $\mathrm{Var}\{\nabla_\theta \ell(\theta^*)\} \approx \nabla_\theta^2 \ell(\hat{\theta})$, we took our estimator to be

$$\widehat{\mathrm{Var}}(\hat{\theta}) = \{\nabla_\theta^2 \ell(\hat{\theta}) + \alpha M\}^{-1} \{\nabla_\theta^2 \ell(\hat{\theta})\} \{\nabla_\theta^2 \ell(\hat{\theta}) + \alpha M\}^{-1}.$$

We lastly evaluated how our estimate for $w(a)$ changed when we changed the number of knots used to define the B-spline basis. Figure S6 shows that the estimate is virtually

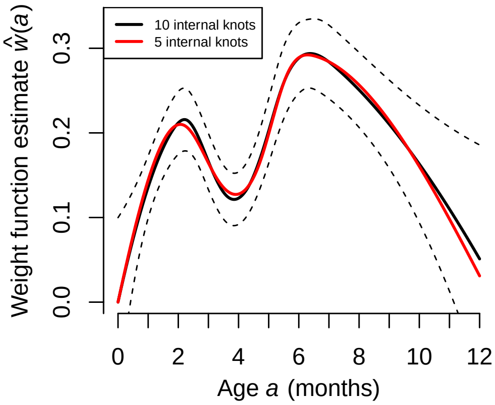


Figure S6: Estimates for $w(a)$ using 10 and 5 internal knots. The dashed black lines give the 95% confidence intervals for the 10 knot estimate.

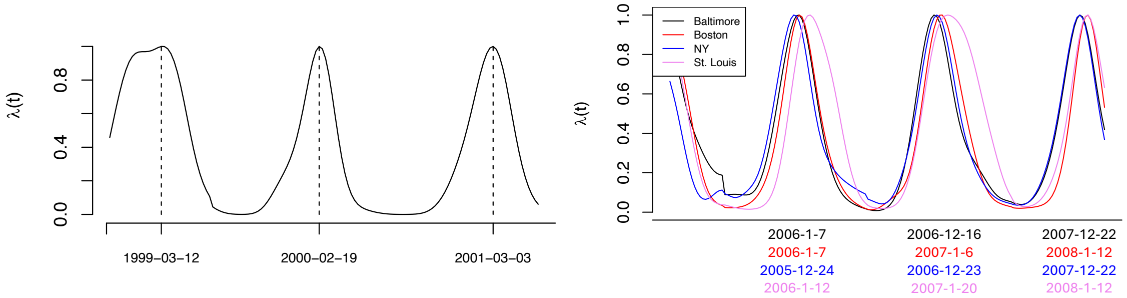


Figure S7: $\lambda(t)$ for COAST (left) and URECA (right) subjects. $\lambda(t)$ is derived from the geographic region covering study subjects and ranges from the earliest birth date to one year after the latest birth date. The dates at the bottom of the plot of $\lambda(t)$ give the peak of RSV season.

identical when we use 10 internal knots (as we did in the main text) and five internal knots.

#### S2.4 Testing our model in COAST and URECA

Values of $\lambda(t)$ covering COAST and URECA subjects are given in Figure S7.

Infection status in each cohort was defined using all data that was available. In URECA, infection status at age one year (365 days) was defined using RSV serology results, as these were the only infection data available in URECA. In COAST, we had first infection age (in days) for 79 out of 285 subjects, RSV serology results in 270 subjects, as well as the age (in days) of the RSV serology blood draw. Since the age of blood draws were not all equal to 365 days, we predicted whether a subject was infected by RSV up until their blood draw age in COAST. Figure S8 shows how we determined RSV infection status and the blood draw age. We treat blood draw ages that exceed 365 days as 365 days because the domain of the weight function did not exceed 365 days. We had $n = 301$ and $n = 242$ subjects from URECA and COAST whose RSV infection status we were able to predict.

We used estimates for $w(a)$ and $\gamma$, $\hat{w}(a)$ and $\hat{\gamma}$, from INSPIRE to calculate the probability a COAST or URECA subject was infected by age $A$. Age $A$ was 365 days for all URECA

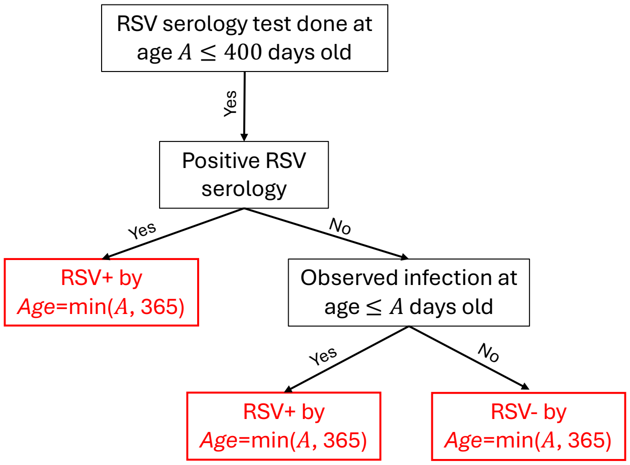


Figure S8: The decision tree used to determine RSV infection status by age "*Age*" in COAST subjects. Red boxes indicate the RSV outcome.

subjects and varied across subjects in COAST. According to Equation 4, the estimated probability for subject $i$ was

$$\hat{\mathbb{P}}(R_i \leq A \mid B_i, z_i) = 1 - \exp\left\{ - \exp(\hat{\gamma}^\top z_i) \int_0^A \lambda(x + B_i)\hat{w}(x)dx \right\}.$$

#### S2.5 Estimating $w(a)$ and $c(a)$ using PRIMA subjects

Birth and hospitalization dates were only available at the granularity of years and months in PRIMA. As such, our observed data was actually an interval spanning one month. For example, if a subject was born in January 2001 and had their first LRTI healthcare encounter in February 2001, we took that to mean the were infected at some point between their first and second months of life. That is, we only knew that $L_i$ lied in the interval $(1 \text{ month}, 2 \text{ months}]$. We fit $w(a)$ and $c(a)$ assuming they were step functions with discontinuities at months $1, \ldots, 11$.

To simplify analyses, we assumed the 12 months were evenly spread out across 365 days. Let $m \in \{1, \ldots, 12\}$ be a month and $k = 365/12$ be the number of days in a month. Under these assumptions, the likelihood for $L_i$ can be expressed as

$$\begin{aligned} \mathbb{P}\{L_i \in (m-1, m] \mid B_i\} =& c(m) \exp\left[ - \int_0^{m-1} \lambda(kx + B_i)\{k\tilde{w}(kx)\}dx \right] \times \\ & \times \left( 1 - \exp\left[ - \int_{m-1}^{m} \lambda(kx + B_i)\{k\tilde{w}(kx)\}dx \right] \right), \end{aligned} \tag{S2}$$

where $\tilde{w}(a)$ is as defined in Methods. The constant $k$ appears as multiplying $\tilde{w}(kx)$ because we have done a change of variables from days to months. To simplify notation, we define

$$\bar{w}(m) = k\tilde{w}(km)$$

to be the weight function expressed as a function of months rather than days. In this notation, $\bar{w}(m)$ is a step function with discontinuities at $m = 1, \ldots, 11$. We also have

$$\mathbb{P}(L_i > 12 \mid B_i) = 1 - \sum_{m=1}^{12} \mathbb{P}\{L_i \in (m-1, m] \mid B_i\}. \tag{S3}$$

To compute the above integrals, we see that because $\bar{w}(\cdot)$ is a step function,

$$\begin{aligned}\int_{m-1}^{m} \lambda(kx + B_i)\bar{w}(x)dx =& \bar{w}(m) \int_{m-1}^{m} \lambda(kx + B_i)dx \\ \int_{0}^{m-1} \lambda(kx + B_i)\bar{w}(x)dx =& \begin{cases} 0 & \text{if } m = 1 \\ \sum_{t=1}^{m-1} \bar{w}(t) \int_{t-1}^{t} \lambda(kx + B_i)dx & \text{if } m > 1 \end{cases},\end{aligned}$$

where we assumed $B_i$ was the first of the month. The corresponding log-likelihood $\ell(\bar{w}, c)$ is a sum of the log-likelihoods for subjects $i$ whose first LRTI healthcare encounter occurred before 12 months (for which we use the likelihood in Equation S2) and whose first encounter occurred after 12 months (for which we use the likelihood in Equation S3).

The estimates for $\bar{w}$ and $c$ were defined to be

$$(\hat{\bar{w}}, \hat{c}) = \underset{\bar{w}, c \in \mathbb{R}^{12}}{\arg\max}\, \ell(\bar{w}, c), \quad 0 \leq \bar{w}, \quad 0 \leq c \leq 1,$$

where we maximized the log-likelihood by iteratively updating $\bar{w}$ and $c$. Updates were derived via constrained Fisher scoring. We finally defined our estimate for $w$ to be $\hat{w} = a\hat{\bar{w}}$, where $a > 0$ was a scalar defined so that the expected number of PRIMA infants infected by age one year matched what was observed in INSPIRE. That is, we set $a$ so that

$$\frac{1}{n}\sum_{i=1}^{n} \hat{\mathbb{P}}(R_i \leq 1 \text{ year} \mid B_i) = 1 - \frac{1}{n}\sum_{i=1}^{n} \exp\left\{ -a \int_{0}^{12} \lambda(kx + B_i)\hat{\bar{w}}(x)dx \right\} = 0.54,$$

where birth months $B_i$ were between June and December (the birth months for INSPIRE subjects) and 0.54 was the observed fraction of INSPIRE infants infected before age 1 year.

#### S2.6 Predicting infection ages in INSPIRE using estimates from PRIMA

Let $\hat{w}(a)$ be the estimate for $w(a)$ derived using PRIMA, where recall age $a$ is in the units of months (see Section S2.5). For INSPIRE subject $i$ with birth date $B_i$, the predicted probability they were infected in month $m \in \{1, \ldots, 12\}$ given they were infected in the first year of life was

$$\begin{aligned}&\hat{\mathbb{P}}\left\{R_i \in (m-1, m] \mid R_i \leq 1 \text{ year}, B_i\right\} \\ =& \frac{\exp\left\{-\int_0^{m-1} \lambda(kx + B_i)\hat{w}(x)dx\right\} - \exp\left\{-\int_0^{m} \lambda(kx + B_i)\hat{w}(x)dx\right\}}{1 - \exp\left\{-\int_0^{12} \lambda(kx + B_i)\hat{w}(x)dx\right\}}.\end{aligned}$$

Here, $\lambda(\cdot)$ is derived from the geographic region covering INSPIRE subjects.

#### S2.7 Estimating the percent infection age variance explained by birth date and non-birth date covariates

We describe our estimator for the fraction of age of first RSV infection variance explained. Our estimator when the outcome was changed to infected by age one year (yes/no) was defined analogously.

Since infection ages were only observed prior to age one year, we do all computations conditional on being infected age one year. Let $R$ be age of first infection, $B$ be birth date, and $z$ be non-birth date covariates. To simplify presentation, we ignore the conditioned event $\{R \leq 1 \text{ year}\}$ in the below derivation. By the tower property of expectation, the variance of $R$ can be expressed as

$$\text{Var}(R) = \text{Var}\left\{\mathbb{E}(R \mid B)\right\} + \mathbb{E}\left[\text{Var}\left\{\mathbb{E}(R \mid B, z) \mid B\right\}\right] + \mathbb{E}\left\{\text{Var}(R \mid B, z)\right\}.$$

The first term is the variance of our prediction for $R$ based solely on birth date. Ignoring the outermost expectation, the second term is interpretable as the variance in our prediction for $R$ based solely on non-birth date covariates, as the variance is being computed while fixing birth date. The outermost expectation simply takes the average of said variance across all birth dates. The third term is the variance that cannot be explained by our model. As such, the percent variance explained by birth date and non-birth date covariates was set to be

$$100 \times \frac{\text{Var}\left\{\mathbb{E}(R \mid B)\right\}}{\text{Var}(R)}, \quad 100 \times \frac{\mathbb{E}\left[\text{Var}\left\{\mathbb{E}(R \mid B, z) \mid B\right\}\right]}{\text{Var}(R)},$$

respectively. We computed these using estimates for $w(a)$ and $\gamma$ derived from INSPIRE, assumed birth dates were uniformly distributed from June 1, 2012 to May 31, 2014 (the period covering INSPIRE births), assumed non-birth date covariates were independent of birth date and distributed as they were in INSPIRE, and used $\lambda(t)$ from the geographic region covering INSPIRE participants.
